# Physician-scientist hiring practices at US universities before and after the COVID-19 pandemic

**DOI:** 10.64898/2026.03.04.26347529

**Authors:** Evan Noch, Aleksandar Obradovic, Snigdha Jain, Jennifer M Kwan

**Affiliations:** Department of Neurology, Division of Neuro-oncology, University of Texas Southwestern Medical Center, Dallas, TX, United States; O’Donnell Brain Institute, University of Texas Southwestern Medical Center, Dallas, TX, United States; Department of Internal Medicine, Columbia University, New York, NY, United States; Yale School of Medicine, Section of Pulmonary Critical Care, New Haven, CT, United States; Yale School of Medicine, Section of Cardiovascular Medicine, New Haven, CT, United States

**Keywords:** Physician-scientist, hiring practices, faculty retention, career development, trainee

## Abstract

Poor retention of physician-scientists in the work force is a major impediment to progress in biomedical research, and the leaky pipeline of junior physician-scientists was exacerbated after the COVID-19 pandemic. We report the results of a multi-institutional survey aimed at assessing hiring practice patterns among academic deans and department chairs, with 34 responses before and 70 responses after the COVID-19 pandemic. We found that private institutions tend to provide greater startup support across all areas of research, including basic science, translational, and clinical arenas, with NIH funding and publication volume predicting the level of support. We found that half of respondents provide research RVUs. The COVID-19 pandemic adversely impacted the availability of supplemental internal funding and bridge funding, which catalyzed institutions to support junior faculty through endowments. Yet, we found that junior faculty had to rearrange clinical schedules to increase clinical productivity. We also found that childcare policies were more robust at private institutions. These data highlight hiring practices across a cohort of academic deans and department chairs to improve transparency of the hiring process for junior faculty candidates approaching their first independent position. Providing greater transparency in hiring practices can help physician-scientist trainees find a good fit for their faculty position and can help stave off attrition from this pipeline.

---

Poor retention of physician-scientists in the work force is a major impediment to progress in biomedical research.^1,2^ Exploring academic hiring practices and their impact from the COVID-19 pandemic can help junior physician-scientists understand the faculty position hiring landscape. Further, shedding light on practices that enable protection of research time and salaries for physician-scientists can be helpful if adapted by institutions trying to retain physician-scientists.

To assess these academic practices, we launched a nationwide survey of academic deans and department chairs at 116 public and private institutions in the US (Figure 1A). The survey was administered via SurveyMonkey (Momentive, San Mateo, California). This study was approved as exempt by the Weill Cornell Medicine Institutional Review. We initially launched the survey in February 2020 immediately before the COVID-19 pandemic. Due to interruptions by the COVID-19 pandemic, we re-launched the survey in August 2022. We received a total of 42 responses pre COVID-19 pandemic, but after filtering for complete responses, a total of 34 responses were included in the analysis (22 public universities, 12 private universities). A total of 108 responses were received post pandemic but after filtering for complete responses, a total of 70 responses were included in the analysis (22 public universities, 48 private universities). Before the COVID-19 pandemic, there were 22 (65%) institutions in the Northeast region, 2 (6%) in the Northwest, 1 (3%) in the South, 7 (21%) in the Southeast, and 2 (6%) in the Southwest (Figure 1B). In the post-pandemic era in August 2022, there were 19 (27%) institutions in the Northeast region, 23 (33%) in the Midwest, 1 (1%) in the Northwest, 2 (3%) in the South, 14 (20%) in the Southeast, 10 (14%) in the Southwest, and 1 (1%) in Alaska/Hawaii. Standard biostatistical analyses were performed, including chi-squared and Fisher’s exact tests, to compare differences between groups. All analyses were performed in R, using R programming language version 4.1.2 (2021–11-01; R Core Team, Vienna, Austria) and RStudio (Integrated Development for R) by RStudio Team (2020; Boston, Massachusetts).

**Figure 1.**
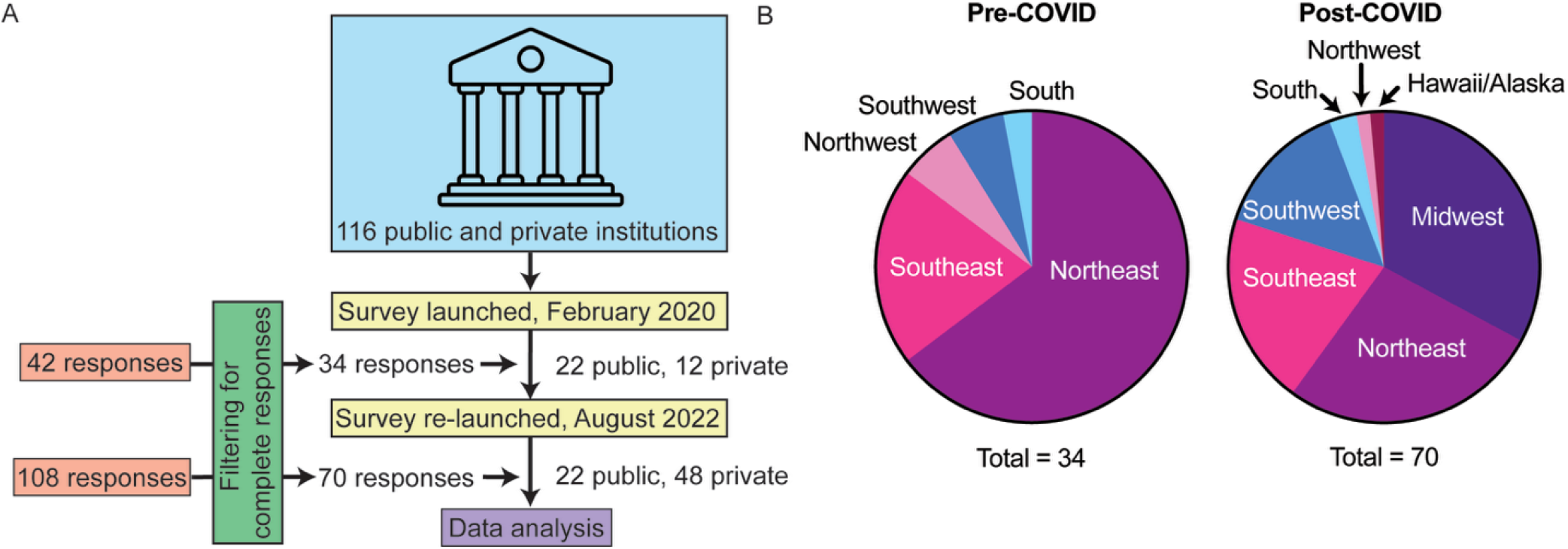
Schema and demographics of survey respondents A. Schematic of survey study and responses. B. Regional demographics of respondents pre- and post-COVID.

When asked about extramural funding for research, about 35% of respondents from public and private institutions felt that applicants should have NIH K or R awards as a pre-requisite for being hired. Private institutions felt that K awards were more important for new faculty hires, whereas public institutions felt that applicants should have R01 awards. More than 40% of public and private institutions reported that they provide bridging support for candidates who are renewing their R01 awards. To monitor the academic success of junior faculty physician-scientists, 43% of respondents reported monitoring NIH funding, and 41% reported monitoring the number of publications.

We found that average startup packages were higher among private than public institutions for candidates in all areas of research, including basic research (mean of $650,000 vs $1,200,000, p= 0.000002), translational research (mean of $344,000 vs $1,020,000, p=0.0195), and clinical research (mean of $339,000 vs $406,000, p=0.026). The most important factors to provide startup funds to junior faculty physician-scientists were NIH funding (28% of respondents) and publications (25% of respondents).

It has been well documented that the COVID-19 pandemic slowed the recruitment of junior faculty physician-scientists.^3-5^ Indeed, we found that 53% of respondents found it more challenging to recruit junior faculty physician-scientists. In the post-COVID era, 73% of respondents said that there was no change in startup packages. However, there were significantly fewer supplemental funds for research activities (p = 0.018). To address this shortcoming, institutions used endowments more commonly to support junior faculty physician-scientists (p = 0.048). Because of the high clinical demand and decreased available research funding post-pandemic, there was a significant increase in junior faculty who had to rearrange their clinical schedule to increase high RVUs (p = 0.00079).

To protect physician-scientist research time and salaries, 50% provided research RVUs, 34% provided funds above the NIH salary cap, 24% distributed salaries equally between clinicians and physician-scientists, 24% of respondents noted they arranged for a higher proportion of time with high RVUs for their physician-scientists, and 21% hired additional clinicians to cover clinical duties (Figure 2A).

**Figure 2.**
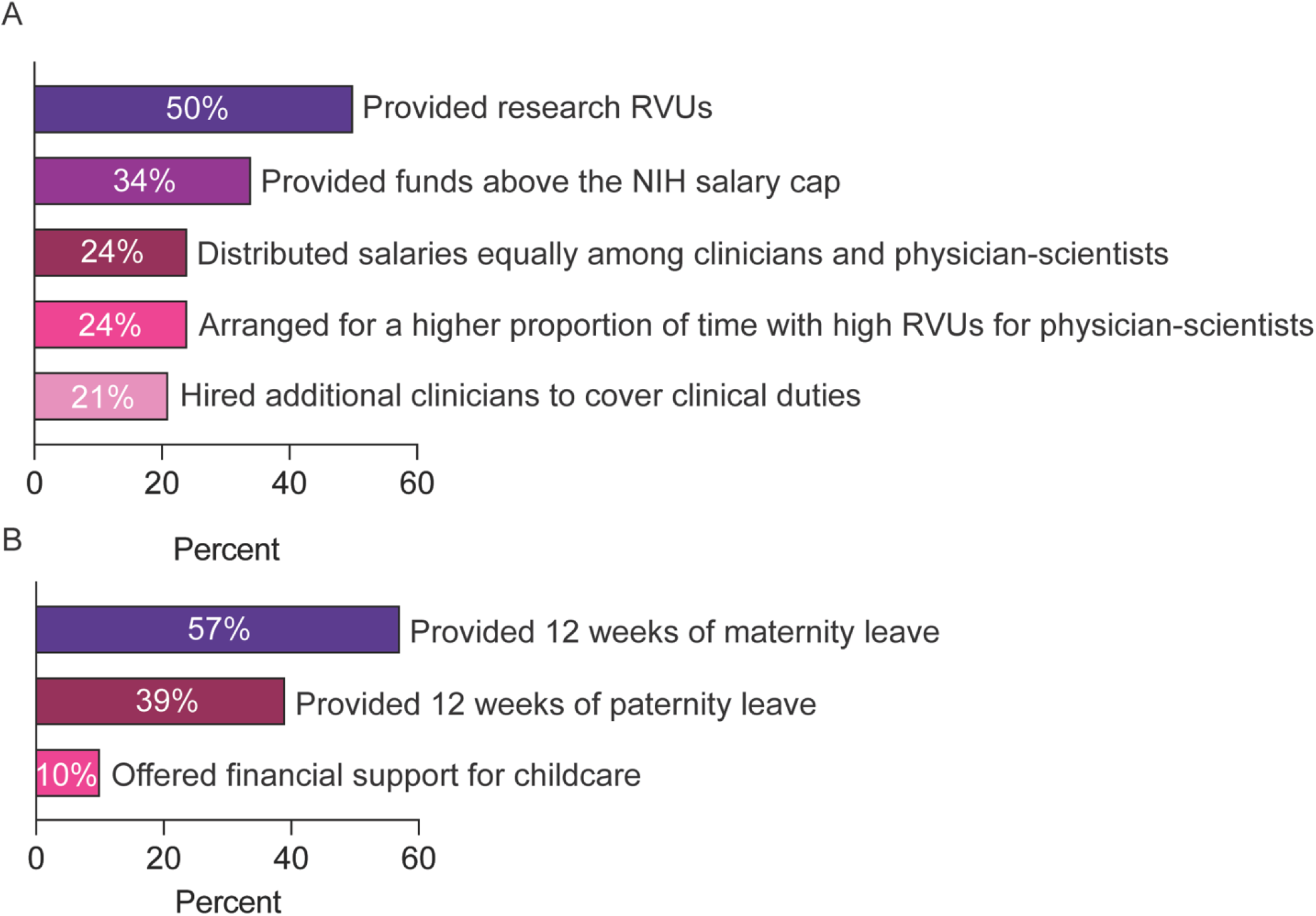
Financial and childcare support of physician-scientists from survey respondents A. Provision of financial assistance to physician-scientists. B. Provision of childcare support to physician-scientists.

We found that 57% and 39% of respondents reported that their institutions provided 12 weeks of maternity and paternity leave, respectively (Figure 2B). 10% offered financial support for childcare, with 56% offering on-campus and 29% offering off-campus childcare. There was also a statistically significant increase in childcare provisions for junior faculty at private versus public institutions.

These results demonstrate that there are distinct hiring practices and support, with significant differences found comparing public versus private institutions, with the latter placing higher value in K award funding and providing higher startup packages for junior faculty physician-scientist recruits. We previously found that female faculty who have young families were particularly affected by the COVID-19 pandemic.^6^ In this study, we found that private institutions provide more childcare support than public institutions, highlighting a key disparity facing recruits with childcare needs. Notably, in terms of research support, we found that 50% of respondents provide research RVUs and 40% support bridge funding during R01 renewal, which are practices aligned with supporting physician-scientists and further exploration of their impact will be important.

As physician-scientist trainees approach their first faculty position, they face a host of challenges, including lack of transparency, the need for extramural funding, and low salary that often preclude them from staying in academia. These can lead to reduced success in physician-scientists obtaining a research faculty position.^7^ Insights on hiring practices can help physician-scientist trainees find a good fit for their faculty position. At the same time, highlighting and increasing adoption of practices that can safeguard research time and balance physician-scientist salaries with that of full-time clinical peers can help stave off attrition from this pipeline.

This study is limited by the small sample size which prevents its generalizability. The study is also limited by the small numbers of responses before the COVID-19 pandemic, preventing full reporting of hiring practices in this era. Future studies will be aimed at a larger sample of institutions and specialties across the US, inclusion of junior faculty experiences during this transition period, and outcomes associated with research support practices.

## Data Availability

All data produced in the present study are available upon reasonable request to the authors

## Acknowledgements

We thank the survey participants for their support of physician-scientists.

